# Continuous theta-burst stimulation of the contralesional primary motor cortex for promotion of upper limb recovery after stroke: a randomized controlled trial

**DOI:** 10.1101/2023.02.28.23286605

**Authors:** Jord JT Vink, Eline CC van Lieshout, Willem M Otte, Ruben PA van Eijk, Mirjam Kouwenhoven, Sebastiaan FW Neggers, H Bart van der Worp, Johanna MA Visser-Meily, Rick M Dijkhuizen

## Abstract

**Background:** Despite improvements in acute stroke therapies and rehabilitation strategies, many stroke patients are left with long-term upper limb motor impairment. We assessed whether an inhibitory repetitive transcranial magnetic stimulation (rTMS) treatment paradigm started within three weeks after stroke onset promotes upper limb motor recovery.

**Methods:** Patients with ischemic stroke or intracerebral hemorrhage and unilateral upper limb motor impairment admitted to a single rehabilitation center were randomized to ten daily sessions of active or sham continuous theta burst stimulation (cTBS) of the contralesional primary motor cortex (M1) combined with standard upper limb therapy, started within three weeks after stroke onset. The primary outcome was the change in the Action Research Arm Test (ARAT) score from baseline (pre-treatment) at three months after stroke. Secondary outcomes included the score on the modified Rankin Scale (mRS) at three months and the length of stay (LOS) at the rehabilitation center. Statistical analyses were performed using mixed models for repeated measures.

**Results:** We enrolled 60 patients between April, 2017 and February, 2021, of whom 29 were randomized to active cTBS and 31 to sham cTBS. One patient randomized to active cTBS withdrew consent before the intervention and was excluded from the analyses. The mean difference in the change in ARAT score from baseline at three months post-stroke was 9.6 points (95%CI 1.2-17.9; p 0.0244) in favor of active cTBS. Active cTBS was associated with better scores on the mRS at three months (OR 0.2; 95%CI 0.1-0.8; p 0.0225) and with an 18 days shorter length of stay at the rehabilitation center than sham cTBS (95%CI 0.0-36.4; p 0.0494). There were no serious adverse events.

**Conclusions:** Ten daily sessions of cTBS of the contralesional M1 combined with upper limb training, started within three weeks after stroke onset, promote recovery of the upper limb, reduce disability and dependence and leads to earlier discharge from the rehabilitation center.

**Trial registration:** The trial was registered at the international clinical trials registry platform (https://trialsearch.who.int/) with unique identifier: NTR6133.

## INTRODUCTION

Upper limb motor impairment is one of the most frequent long-term neurological consequences of ischemic stroke or intracerebral haemorrhage.^1,2^ Despite recent developments in acute stroke treatment and rehabilitative therapy, recovery of upper limb motor function is often incomplete, resulting in limitations in functioning and participation.^3–6^ More effective rehabilitation strategies that improve stroke recovery and lead to better clinical outcomes are therefore required.^7^

Spontaneous recovery after stroke is believed to be driven by neurobiological processes that occur mainly during a period of heightened brain plasticity in the first three months after stroke.^8,9^ Conventional rehabilitation strategies focus on regaining function within this period primarily through physical therapy. Previous studies have shown that patients who have recovered from stroke present a more symmetrical inhibitory drive between the primary motor cortices.^10–14^ An increased and persistent inhibitory drive from the contralesional to the ipsilesional primary motor cortex (M1) has been associated with more severe post-stroke motor deficits.^10–13^ It has been suggested that restoration of the interhemispheric balance can result in a brain state that is more prone to spontaneous recovery and rehabilitation therapy of the affected arm.^13^ On the other hand, excitability of the contralesional M1 may be within normal levels after stroke^15^ and others have suggested that an interhemispheric imbalance might be a consequence of underlying processes of motor recovery.^15,16^

An interhemispheric imbalance may be restored through excitation of the lesioned M1 or inhibition of the contralesional M1.^17^ Repetitive transcranial magnetic stimulation (rTMS) is a non-invasive brain stimulation method to up-^18^ or downregulate^19^ cortical excitability. Inhibitory rTMS of the contralesional M1 combined with upper limb therapy was initially investigated as a treatment for the promotion of upper limb recovery in chronic stroke patients.^20^ However, a recent guidelines paper and a meta-analysis of randomized controlled trials indicate that inhibition of the contralesional M1 using inhibitory low-frequency (LF) rTMS followed by upper limb therapy is more effective in promoting upper limb recovery when started within the first two to three months post-stroke.^21,22^ This has been shown to be the sensitive period for improvement of motor recovery^23^, presumably associated with heightened plasticity, which could potentially also facilitate a therapeutic effect of rTMS. Yet, it remains to be determined whether additional recovery achieved with rTMS treatment reduces disability and dependency and whether the effects persist after the first three months post-stroke.

Conventional LF rTMS treatment consists of 15-minute sessions, while continuous theta-burst stimulation (cTBS), a novel inhibitory rTMS paradigm, has a much shorter treatment duration of 40 seconds per session. Therefore, the use of cTBS can improve patient comfort and increase cost-effectiveness of the intervention.^24,25^ So far, the effect of cTBS on upper limb recovery has only been tested in a single randomized trial randomizing only 14 patients to receive active cTBS group and 13 patients to receive sham cTBS.^26^ This did not reveal an effect of cTBS started within 10 weeks after stroke on the change on a multifaceted upper limb motor function score (obtained from four upper limb function tests) within 30 days after treatment with respect to the change in this score (over a period of 7 days) before treatment. We assessed whether two weeks of daily contralesional cTBS started within the first three weeks after stroke improves long-term upper limb motor recovery up to 12 months post-stroke.

## METHODS

### Study design

We performed a single-center, prospective, randomized, sham-controlled clinical trial with a single-blind intervention and a double-blind primary outcome evaluation at rehabilitation center De Hoogstraat (Utrecht, The Netherlands), according to CONSORT guidelines. A summary of the trial protocol has been published^27^ and the full protocol is available in the supplementary materials. The study was approved by the Medical Research Ethics Committee of the University Medical Center Utrecht.

### Participants

We included patients aged 18 years or older with first-ever ischemic stroke or intracerebral hemorrhage and a paresis of one arm, as defined by a Motricity Index (MI)^28^ between 9 and 99, in whom treatment could be started within three weeks after stroke onset.^29^ Patients were excluded from participation if they had another disabling medical condition, as determined by the treating physician; could use the hand of the paretic arm (almost) normally (MI pinch grip score of 33); had a severe deficit in communication, memory, or understanding that would impede proper study participation; or had a contraindication to rTMS according to TMS safety guidelines.^30^ All patients gave written informed consent.

### Randomization and masking

Patients were randomly assigned to ten daily sessions of contralesional cTBS or to sham cTBS during two weeks, in addition to regular care upper limb therapy, using a secured online allocation system (Research Online V2.0, Julius Centre, the Netherlands) by the investigator performing the treatment. Randomization was stratified according to the ability to extend one or more fingers of the paretic arm.^31^ Sham cTBS was performed at 10% of the resting motor threshold (RMT), which was defined as the minimum machine output (MO) at which stimulation evoked at least five out of ten MEPs with a peak-to-peak amplitude of over 50 μV^25^, with the TMS coil rotated 45 degrees relative to the scalp, and patients were masked to treatment allocation using auditory masking of the TMS coil sound.

### Procedures

Treatment was delivered in ten daily sessions on consecutive working days. cTBS was delivered over the contralesional M1, which was defined as the position on the scalp at which motor-evoked potentials (MEPs) with the largest peak-to-peak amplitude could be evoked in the contralateral first dorsal interosseous (FDI) muscle by delivering TMS pulses and monitoring the electromyogram (EMG). A Neuro-MS/D advanced therapeutic magnetic stimulator and an angulated 100 mm figure-of-eight TMS coil (Neurosoft, Ivanovo, Russia) were used for stimulation. EMG was recorded, amplified, and digitized at a sampling frequency of 20 kHz using a four-channel Neuro-MEP amplifier (Neurosoft, Ivanovo, Russia). cTBS consisted of continuous delivery of three stimuli bursts at 50 Hz repeated at five bursts per second for a duration of forty seconds with a biphasic TMS-induced current at 45 degrees to the midline. Stimulation intensity was set at 70% of the RMT. TMS coil placement was recorded using a neuronavigation system (Brain Science Tools BV, De Bilt, The Netherlands) starting from the 16^th^ patient. cTBS or sham cTBS was delivered 15 minutes before standard upper limb therapy, which consisted of a 60-minute group therapy session of individualized upper limb exercises, according to the Concise Arm and hand Rehabilitation Approach in Stroke (CARAS).^32^ CARAS consists of a daily training program during which patients perform specific exercises with the paretic arm under guidance of physical or occupational therapists, supplemented with exercises they can perform independently during the rest of the day.

### Outcomes

The primary outcome measure was the change in the action research arm test (ARAT) score from baseline at three months after stroke, as recommended by the Stroke Recovery and Rehabilitation Roundtable (SRRR).^33^ The ARAT is a performance test that assesses the ability to perform gross movements and to grasp, move and release objects differing in size, weight, and shape, of which validity and reliability have been demonstrated previously.^34,35^ The ARAT ranges from 0 to 57, with higher scores indicating better performance. The ARAT was measured at three months after stroke by an independent trained physician assistant, blinded to treatment allocation.

Predefined secondary outcomes included the change in ARAT score <12 hours, 1 week and 1 month post-treatment, and 6 and 12 months post-stroke. Other predefined secondary outcomes were tests that assess different domains of the international classification of functioning, disability, and health (ICF) framework.^36^ These included the upper limb section of the Fugl-Meyer assessment (FMA) score^37^ for motor impairment; the stroke upper limb capacity score (SULCS),^38^ Jebsen Taylor test (JTT)^39^ score, Barthel Index^40^ and Nine Hole Peg Test (NHPT)^41^ for activity; the modified Rankin Scale (mRS)^42^ for disability; and the upper limb section of the stroke impact scale (SIS-UL)^43^ and EuroQol(EQ)-5D^44^ for quality of life. Secondary outcomes were assessed by an investigator who was aware of the treatment allocation. Table 2 shows an overview of the measures that were assessed for each visit. We also assessed the length of stay (LOS) in the rehabilitation center, the dose of upper limb therapy and self-practice during the two-week treatment period and the week thereafter, and the contralesional RMT before each cTBS session. We monitored and recorded (serious) adverse events (AEs) that occurred during the two-week treatment period or one week thereafter.

**Table 1.**
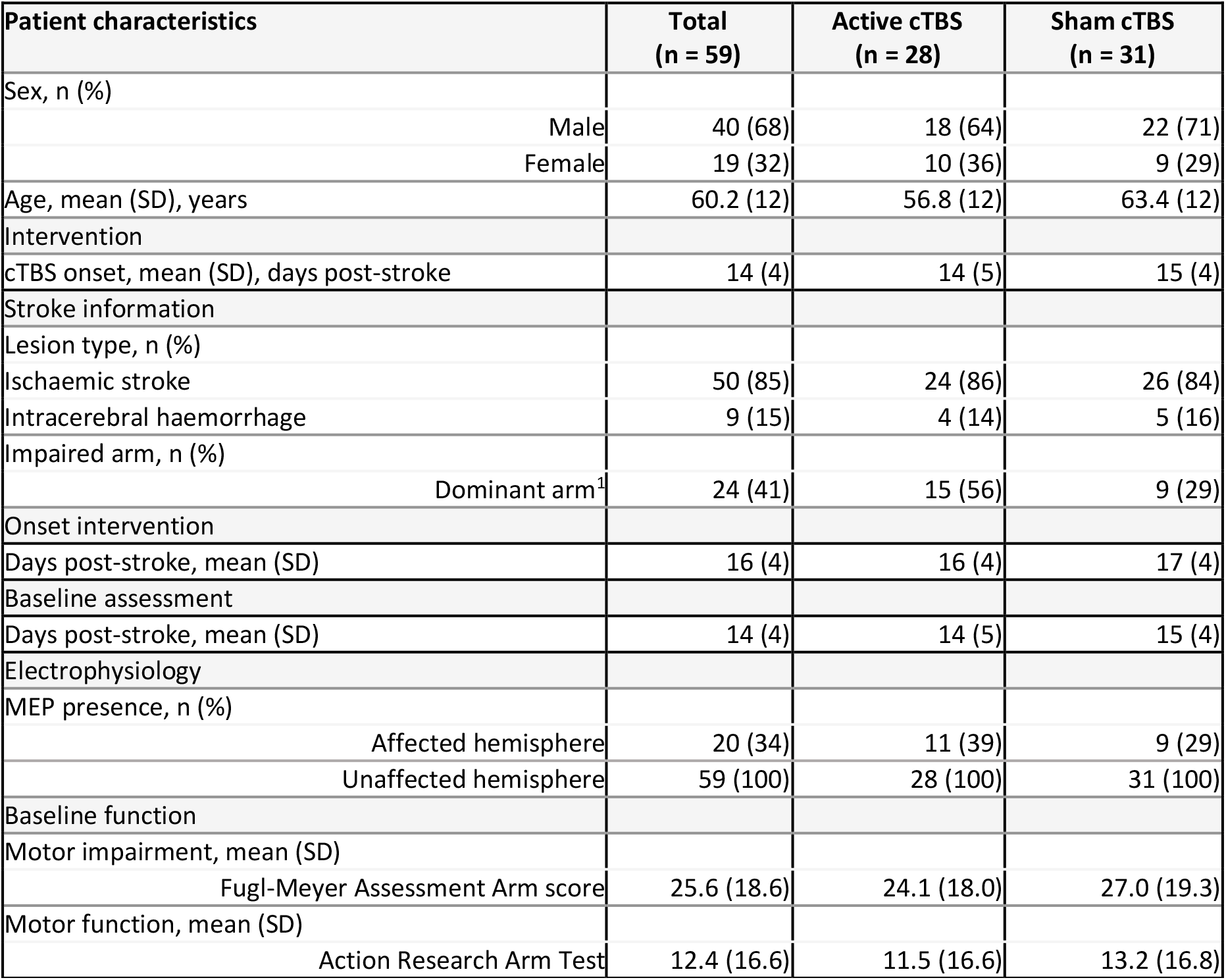
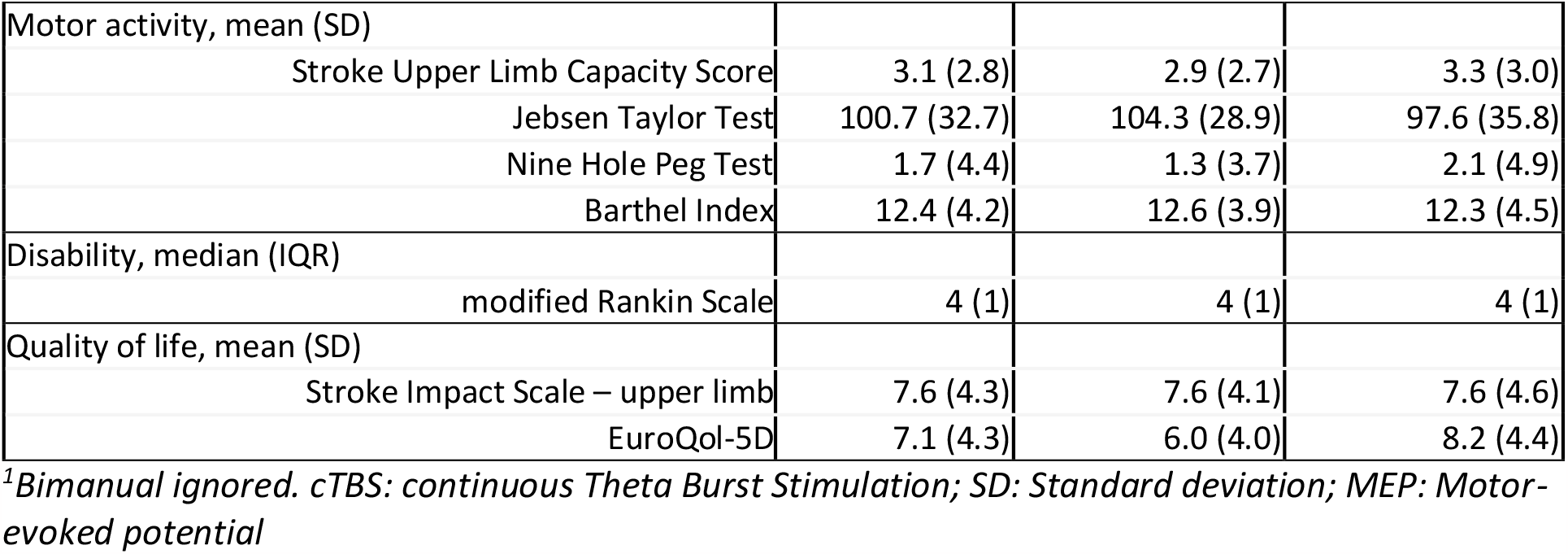
Baseline characteristics (full table available in online supplement)

| Patient characteristics | Total<br>(n = 59) | Active cTBS<br>(n = 28) | Sham cTBS<br>(n = 31) |
| --- | --- | --- | --- |
| Sex, n (%) |  |  |  |
| Male | 40 (68) | 18 (64) | 22 (71) |
| Female | 19 (32) | 10 (36) | 9 (29) |
| Age, mean (SD), years | 60.2 (12) | 56.8 (12) | 63.4 (12) |
| Intervention |  |  |  |
| cTBS onset, mean (SD), days post-stroke | 14 (4) | 14 (5) | 15 (4) |
| Stroke information |  |  |  |
| Lesion type, n (%) |  |  |  |
| Ischaemic stroke | 50 (85) | 24 (86) | 26 (84) |
| Intracerebral haemorrhage | 9 (15) | 4 (14) | 5 (16) |
| Impaired arm, n (%) |  |  |  |
| Dominant arm<sup>1</sup> | 24 (41) | 15 (56) | 9 (29) |
| Onset intervention |  |  |  |
| Days post-stroke, mean (SD) | 16 (4) | 16 (4) | 17 (4) |
| Baseline assessment |  |  |  |
| Days post-stroke, mean (SD) | 14 (4) | 14 (5) | 15 (4) |
| Electrophysiology |  |  |  |
| MEP presence, n (%) |  |  |  |
| Affected hemisphere | 20 (34) | 11 (39) | 9 (29) |
| Unaffected hemisphere | 59 (100) | 28 (100) | 31 (100) |
| Baseline function |  |  |  |
| Motor impairment, mean (SD) |  |  |  |
| Fugl-Meyer Assessment Arm score | 25.6 (18.6) | 24.1 (18.0) | 27.0 (19.3) |
| Motor function, mean (SD) |  |  |  |
| Action Research Arm Test | 12.4 (16.6) | 11.5 (16.6) | 13.2 (16.8) |
| Motor activity, mean (SD) |  |  |  |
| Stroke Upper Limb Capacity Score | 3.1 (2.8) | 2.9 (2.7) | 3.3 (3.0) |
| Jebsen Taylor Test | 100.7 (32.7) | 104.3 (28.9) | 97.6 (35.8) |
| Nine Hole Peg Test | 1.7 (4.4) | 1.3 (3.7) | 2.1 (4.9) |
| Barthel Index | 12.4 (4.2) | 12.6 (3.9) | 12.3 (4.5) |
| Disability, median (IQR) |  |  |  |
| modified Rankin Scale | 4 (1) | 4 (1) | 4 (1) |
| Quality of life, mean (SD) |  |  |  |
| Stroke Impact Scale – upper limb | 7.6 (4.3) | 7.6 (4.1) | 7.6 (4.6) |
| EuroQol-5D | 7.1 (4.3) | 6.0 (4.0) | 8.2 (4.4) |

<sup>1</sup>*Bimanual ignored. cTBS: continuous Theta Burst Stimulation; SD: Standard deviation; MEP: Motor-evoked potential*

**Table 2.**
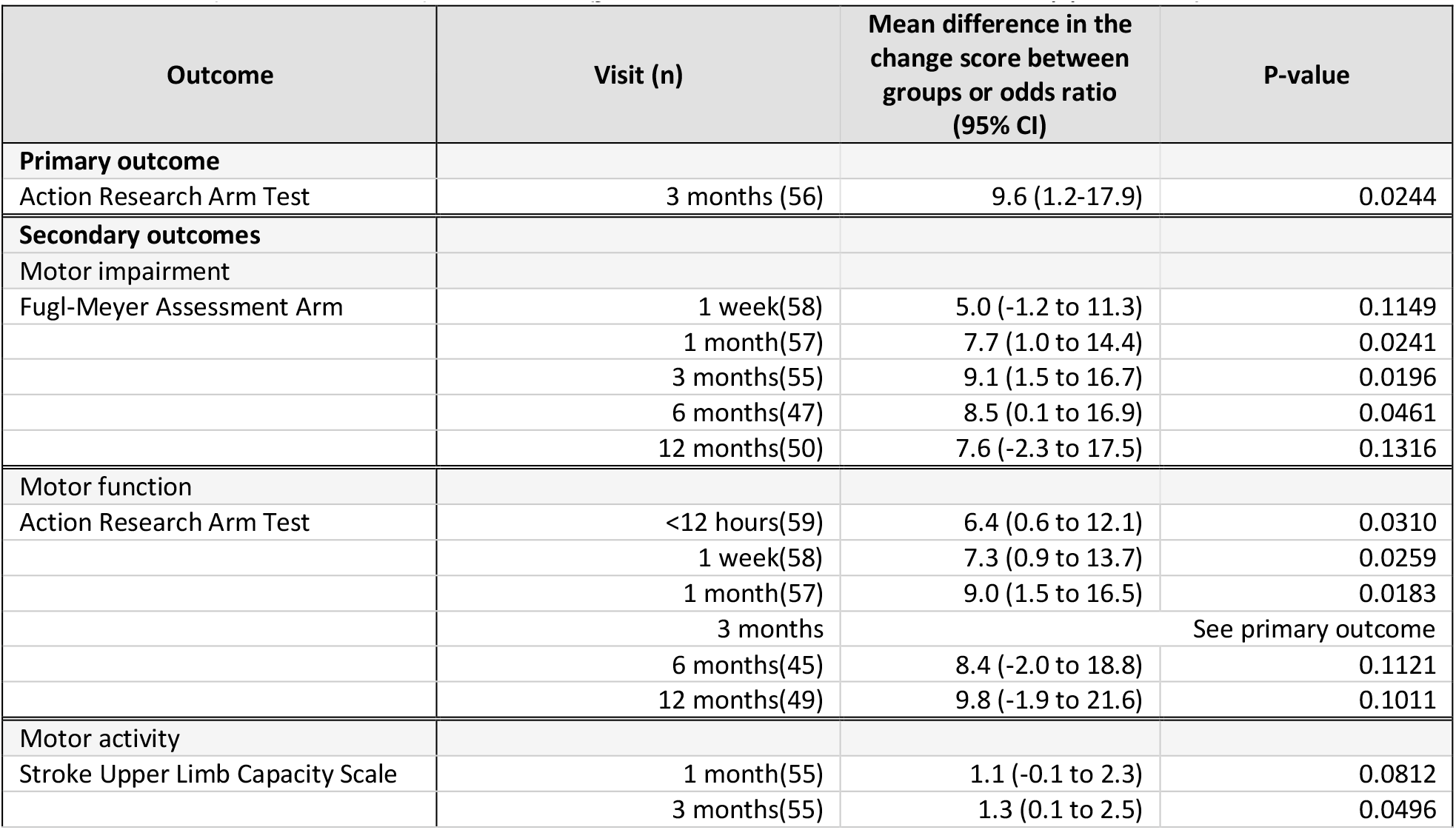
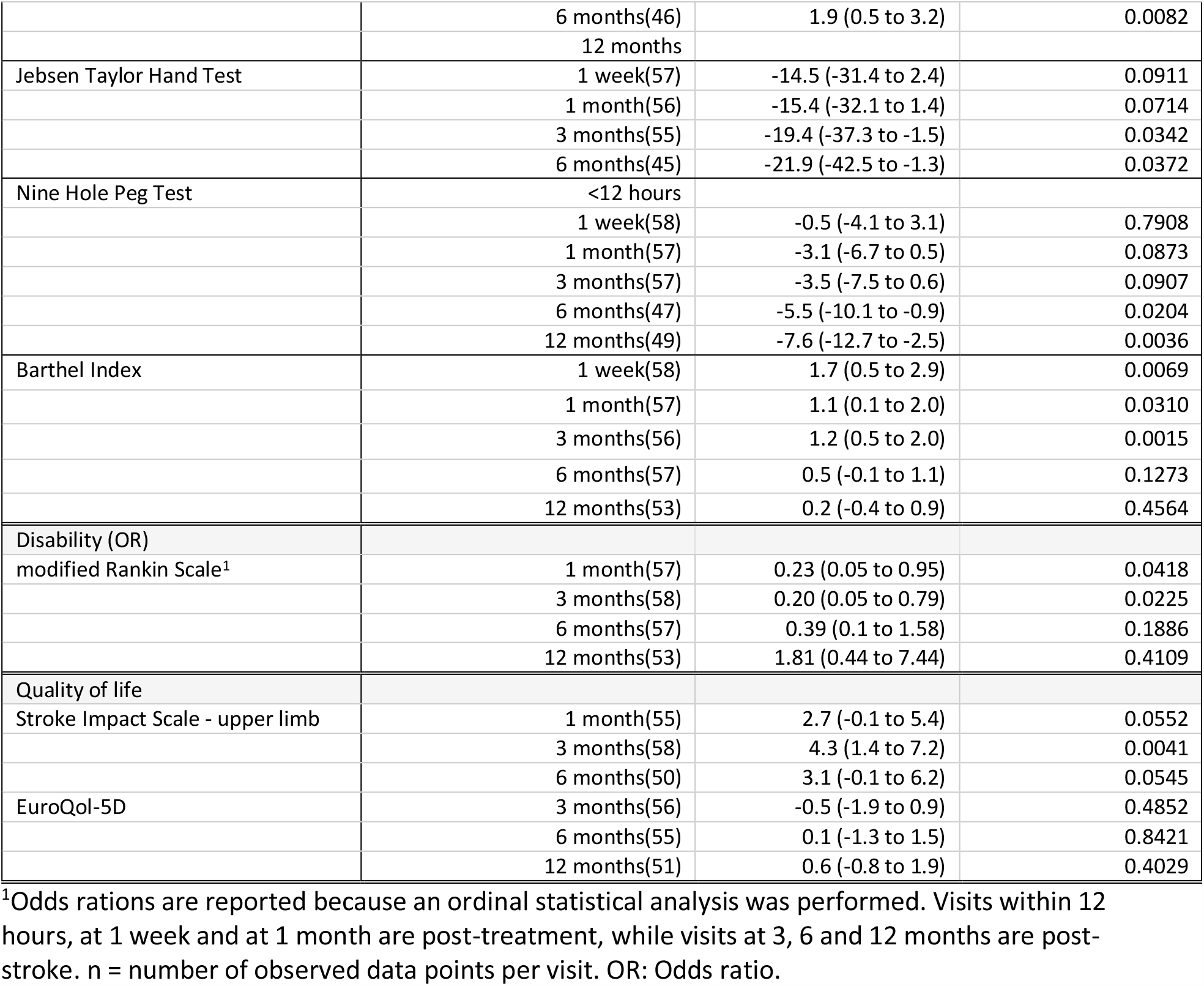
Primary and secondary outcomes (full table available in online supplement).

| Outcome | Visit (n) | Mean difference in the change score between groups or odds ratio (95% CI) | P-value |
| --- | --- | --- | --- |
| **Primary outcome** |  |  |  |
| Action Research Arm Test | 3 months (56) | 9.6 (1.2-17.9) | 0.0244 |
| **Secondary outcomes** |  |  |  |
| **Motor impairment** |  |  |  |
| Fugl-Meyer Assessment Arm | 1 week(58) | 5.0 (-1.2 to 11.3) | 0.1149 |
|  | 1 month(57) | 7.7 (1.0 to 14.4) | 0.0241 |
|  | 3 months(55) | 9.1 (1.5 to 16.7) | 0.0196 |
|  | 6 months(47) | 8.5 (0.1 to 16.9) | 0.0461 |
|  | 12 months(50) | 7.6 (-2.3 to 17.5) | 0.1316 |
| **Motor function** |  |  |  |
| Action Research Arm Test | <12 hours(59) | 6.4 (0.6 to 12.1) | 0.0310 |
|  | 1 week(58) | 7.3 (0.9 to 13.7) | 0.0259 |
|  | 1 month(57) | 9.0 (1.5 to 16.5) | 0.0183 |
|  | 3 months | See primary outcome |  |
|  | 6 months(45) | 8.4 (-2.0 to 18.8) | 0.1121 |
|  | 12 months(49) | 9.8 (-1.9 to 21.6) | 0.1011 |
| **Motor activity** |  |  |  |
| Stroke Upper Limb Capacity Scale | 1 month(55) | 1.1 (-0.1 to 2.3) | 0.0812 |
|  | 3 months(55) | 1.3 (0.1 to 2.5) | 0.0496 |
|  | 6 months(46) | 1.9 (0.5 to 3.2) | 0.0082 |
|  | 12 months |  |  |
| Jebsen Taylor Hand Test | 1 week(57) | -14.5 (-31.4 to 2.4) | 0.0911 |
|  | 1 month(56) | -15.4 (-32.1 to 1.4) | 0.0714 |
|  | 3 months(55) | -19.4 (-37.3 to -1.5) | 0.0342 |
|  | 6 months(45) | -21.9 (-42.5 to -1.3) | 0.0372 |
| Nine Hole Peg Test | <12 hours |  |  |
|  | 1 week(58) | -0.5 (-4.1 to 3.1) | 0.7908 |
|  | 1 month(57) | -3.1 (-6.7 to 0.5) | 0.0873 |
|  | 3 months(57) | -3.5 (-7.5 to 0.6) | 0.0907 |
|  | 6 months(47) | -5.5 (-10.1 to -0.9) | 0.0204 |
|  | 12 months(49) | -7.6 (-12.7 to -2.5) | 0.0036 |
| Barthel Index | 1 week(58) | 1.7 (0.5 to 2.9) | 0.0069 |
|  | 1 month(57) | 1.1 (0.1 to 2.0) | 0.0310 |
|  | 3 months(56) | 1.2 (0.5 to 2.0) | 0.0015 |
|  | 6 months(57) | 0.5 (-0.1 to 1.1) | 0.1273 |
|  | 12 months(53) | 0.2 (-0.4 to 0.9) | 0.4564 |
| Disability (OR) |  |  |  |
| modified Rankin Scale<sup>1</sup> | 1 month(57) | 0.23 (0.05 to 0.95) | 0.0418 |
|  | 3 months(58) | 0.20 (0.05 to 0.79) | 0.0225 |
|  | 6 months(57) | 0.39 (0.1 to 1.58) | 0.1886 |
|  | 12 months(53) | 1.81 (0.44 to 7.44) | 0.4109 |
| Quality of life |  |  |  |
| Stroke Impact Scale - upper limb | 1 month(55) | 2.7 (-0.1 to 5.4) | 0.0552 |
|  | 3 months(58) | 4.3 (1.4 to 7.2) | 0.0041 |
|  | 6 months(50) | 3.1 (-0.1 to 6.2) | 0.0545 |
| EuroQol-5D | 3 months(56) | -0.5 (-1.9 to 0.9) | 0.4852 |
|  | 6 months(55) | 0.1 (-1.3 to 1.5) | 0.8421 |
|  | 12 months(51) | 0.6 (-0.8 to 1.9) | 0.4029 |

<sup>1</sup>Odds ratios are reported because an ordinal statistical analysis was performed. Visits within 12 hours, at 1 week and at 1 month are post-treatment, while visits at 3, 6 and 12 months are post-stroke. n = number of observed data points per visit. OR: Odds ratio.

### Statistical analysis

The sample size calculation was based on an effect size of 0.55 on the ARAT score obtained from a meta-analysis.^27^ To detect the hypothesized effect with 80% power, and a two-sided alpha of 0.05, a total sample size of 56 patients was required. We included sixty patients, with thirty patients per group, to account for loss to follow-up.

We followed the pre-specified statistical analysis plan, which is available in the supplementary materials and was completed prior to data-lock. The primary outcome was the change in ARAT score from baseline (pre-treatment) at three months post-stroke.^27^ The primary analysis was performed using a linear mixed model for repeated measures (MMRM) with an unstructured variance-covariance matrix in the intention-to-treat (ITT) population, includes all randomized participants irrespective of follow-up. Missing data were assumed to be missing at random. The model included the baseline value of the investigated outcome; the stratification factor (ability vs. no ability to extend one or more fingers), visit (<12 hours, 1 week and 1 month post-treatment, and 3, 6 and 12 months post-stroke) and the interaction of treatment (sham cTBS; active cTBS) by visit. We performed a sensitivity analysis of the primary outcome in the per-protocol (PP) population (defined as those who had an ARAT score assessed at three months) and additional sensitivity analyses in which additional covariates, i.e. MEP status and dominant hand paresis, were included in the main analysis.

We performed similar analyses for the effect of treatment on the secondary outcomes at all visits except for the mRS and LOS. The mRS was analyzed using a cumulative link mixed model of the total mRS scores due to the ordinal nature of the data. The effect of treatment on the LOS and the dose of upper limb therapy and self-practice that patients received were analyzed with independent samples t-tests. The effect of treatment on excitability of the contralesional M1 was analyzed using a linear mixed-effects model (LME). The outcome was the contralesional RMT determined before each cTBS session and the LME included the baseline RMT, number of cTBS session (1 to 10) and type of treatment (sham cTBS; active cTBS). Pearson’s correlation was used to calculate the correlation between the change in contralesional RMT during the treatment period and the change in ARAT score between baseline and 3 months post-stroke.

Statistical analysis was performed with R 4.1 and SPSS 26.0 (IBM, Chicago, IL, USA). All hypotheses were tested a two-sided alpha of 0.05.

## RESULTS

Between April 14, 2017 and February 12, 2021, 494 stroke patients with arm weakness were screened for eligibility, of whom 60 were enrolled (Figure 1). Twenty-nine patients were randomly assigned to receive active cTBS, of whom one withdrew consent before starting treatment, leaving 28 patients in the cTBS group, and 31 patients were assigned to receive sham cTBS. Therefore, the ITT population consisted of 59 patients. The ARAT could not be assessed at three months in three patients (one in the active cTBS group and two in the sham cTBS group) due to physical limitations (shoulder pain, tiredness) and one patient in the sham cTBS group had a recurrent stroke. These four patients were excluded from the PP population (Figure 1).

**Figure 1.**
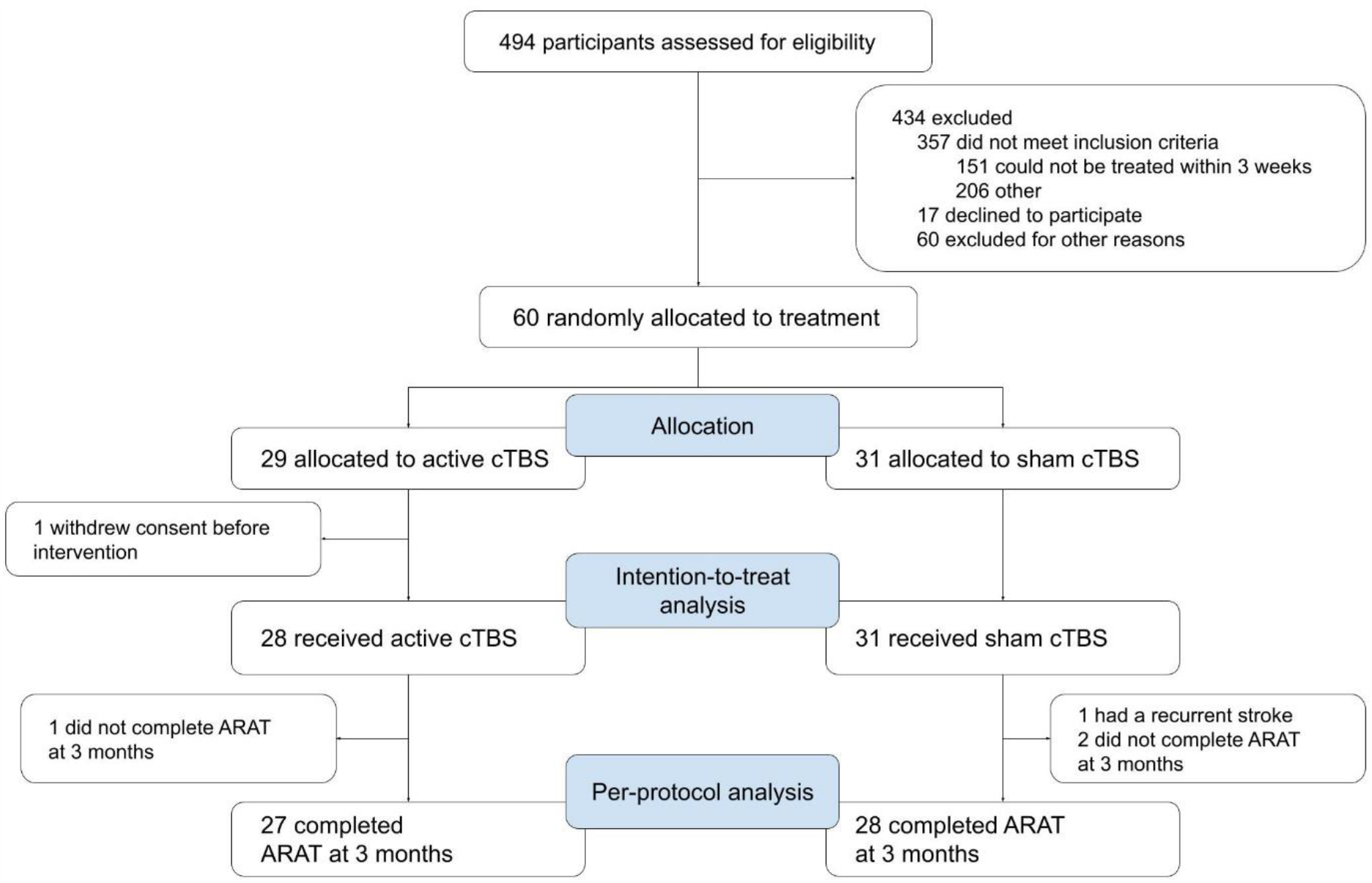
Participant flow diagram

Patient characteristics are shown in Table 1. None of the patients had previously been treated with cTBS.

The <12 hours, 1 week and 1 month post-treatment visits were assessed 29 (SD 4), 36 (SD 5) and 59 (SD 7) days post-stroke, respectively, and the 3, 6 and 12 months post-stroke visits were assessed 90 (SD 4), 184 (SD 11) and 368 (SD 10) days post-stroke, respectively.

The change in ARAT score from baseline at three months post-stroke was 27.6 points in the active cTBS group compared to 18.0 points in the sham cTBS group, with a mean difference of 9.6 points (95% CI 1.2 to 17.9; p 0.0244; Figure 2A). Sensitivity analysis showed a mean difference in ARAT change score of 9.9 points (95% CI 1.1 to 18.7; p 0.0276) in the PP population in favor of active cTBS. Sensitivity analyses including baseline MEP status or dominant hand paresis as additional covariate showed mean differences in ARAT change score of 8.4 points (95% CI 0.3 to 16.5; p 0.0414) and 9.2 points (95% CI 0.7 to 17.6; p 0.0341), respectively, both in favor of active cTBS.

**Figure 2.**
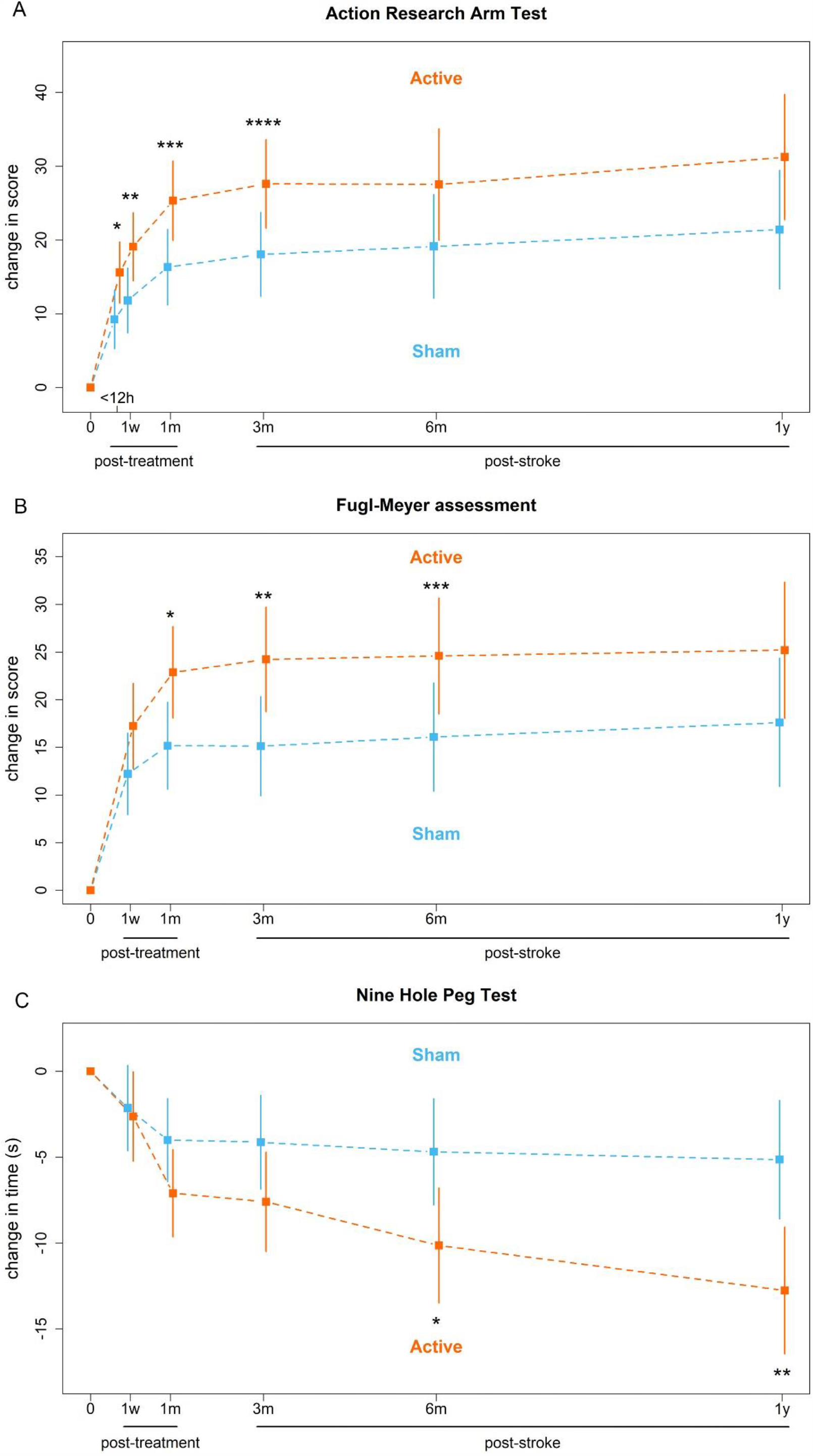
Mean and 95% confidence intervals of the changes in (**A**) Action Research Arm Test (ARAT) scores, (**B**) Fugl-Meyer assessment (FMA) arm scores and (**C**) Nine Hole Peg Test (NHPT) times for the active and sham cTBS groups calculated using mixed-effects model for repeated measures. 0: baseline; h: hours; w: weeks; m: months; y: years (**A**) ARAT score ranges from 0 to 57 points and a higher score indicates better outcome. *p 0.0310. **p 0.0259. ***p 0.0183. ****p 0.0244. (**B**) FMA score ranges from 0 to 66 points and a higher score indicates better outcome. *p 0.0241. **p 0.0196. ***p 0.0461. (**C**). Maximum NHPT time is 50 seconds and a lower time indicates better outcome. *p 0.0204. **p 0.0036.

In the secondary analyses, the mean difference at three months post-stroke between active and sham cTBS groups was 9.1 points on the FMA (95% CI 1.5 to 16.7; p 0.0196; Figure 2B), 1.3 points on the SULCS (95% CI 0.1 to 2.5; p 0.0496), -19.4 points on the JTT (95% CI - 37.3 to -1.5; p 0.0342), -3.5 points on the NHPT (95% CI -7.5 to 0.6; p 0.0907; Figure 2C), 1.2 points on the BI (95% CI 0.5 to 2.0; p 0.0015), 4.3 points on the SIS-UL (95% CI 1.4 to 7.2; p 0.0041) and -0.5 on the EQ-5D (95% CI -1.9 to 0.9; p 0.4852), all indicating better outcomes except for the NHPT and EQ-5D. Score on the mRS were better with active than with sham cTBS (odd ratio 0.2; 95% CI 0.1 to 0.8; p 0.0225; Figure 3). Details are presented in Table 2.

**Figure 3.**
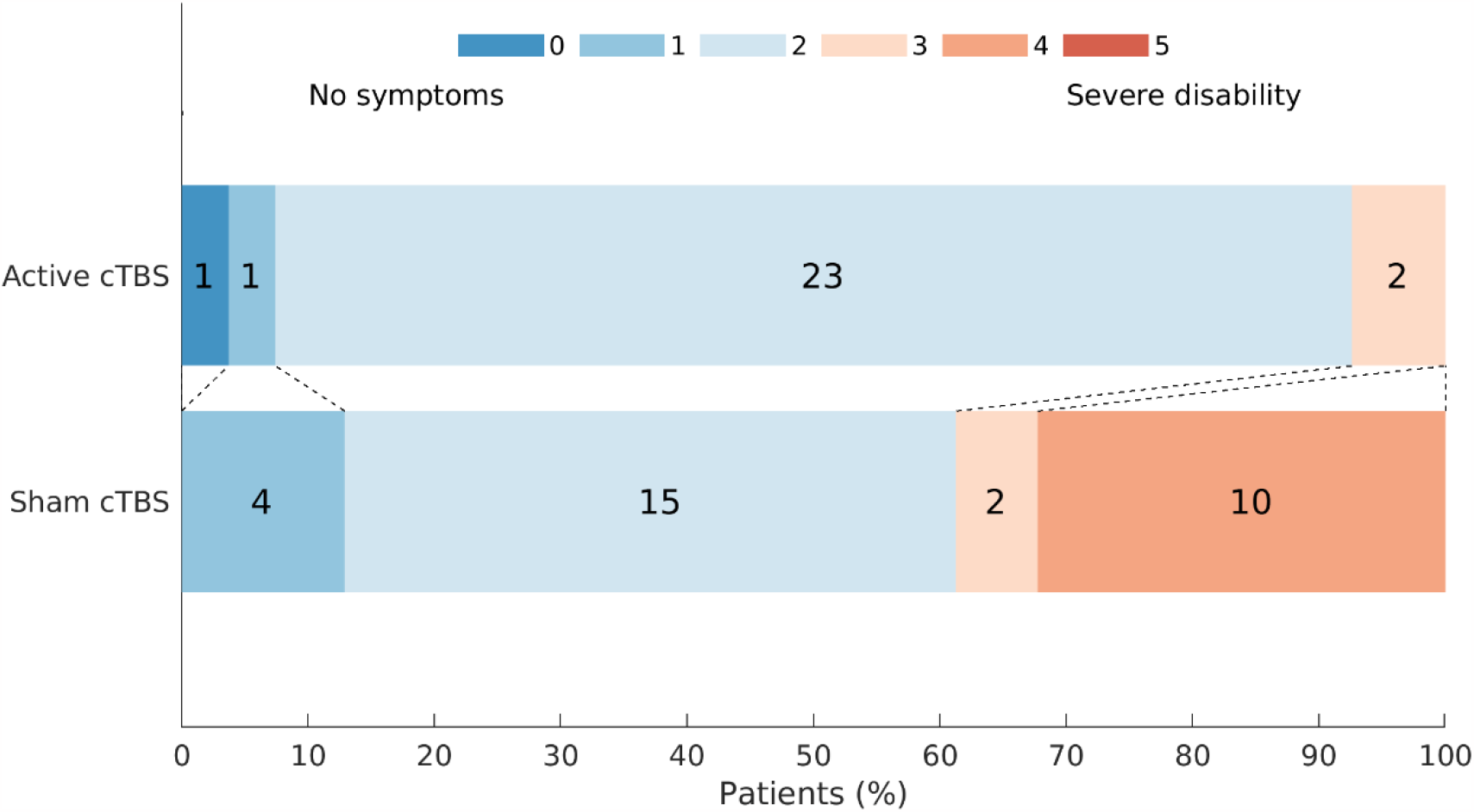
Modified Rankin Scale (mRS) scores at three months post-stroke for the active and sham cTBS groups. The mRS ranges from 0 to 5 and a lower score indicates better outcome. p 0.0225.

The mean LOS was 63 days in the active cTBS group compared to 81 days in the sham cTBS group with a mean difference of 18 days (95% CI 0.0 to 36.4; p 0.0494). Mean duration of upper limb therapy in the first three weeks after TMS treatment onset was 17.0 hours in the active cTBS group compared to 17.4 hours in the sham cTBS group (95% CI -2.2 to 1.4; p 0.6628). Mean duration of independent self-practice was 8.1 hours in the active cTBS group compared to 8.5 hours of in the sham cTBS group (95% CI -4.9 to 4.1; p 0.8572). Active cTBS resulted in a mean increase of 1.4% of the contralesional RMT (95% CI -0.0 to 2.7; p 0.0515). The change in contralesional RMT during the treatment was not associated with the change in ARAT score between baseline and 3 months post-stroke in the active cTBS group (r 0.02; p 0.9082).

No serious adverse events (SAEs) were reported. Headache was the most prevalent side effect, which occurred more frequently with active cTBS (in ten out of 279 sessions) than with sham cTBS (in three out of 307 sessions). Other side effects (i.e., muscle pain and nausea) were rare (Table 3).

**Table 3.** Adverse events reported during the two-week treatment period and 1 week thereafter.

| Adverse event, n (%) | Total (n=586<sup>1</sup>) | Active cTBS (n=279) | Sham cTBS (n=307) | P-value |
| --- | --- | --- | --- | --- |
| Nausea | 1 (0.17) | 0 (0) | 1 (0.33) | 0.524 |
| Headache | 13 (2.22) | 10 (3.58) | 3 (0.98) | 0.030 |
| Muscle pain | 2 (0.34) | 2 (0.72) | 0 (0) | 0.226 |
| Sensory impairment | 1 (0.17) | 0 (0) | 1 (0.33) | 0.524 |
| Slowed thinking | 2 (0.34) | 0 (0) | 2 (0.65) | 0.274 |

<sup>1</sup>Total amount of cTBS sessions performed.

## DISCUSSION

Patients who received active cTBS showed greater improvement in upper limb recovery as measured with the ARAT at three months post-stroke compared to patients who received sham cTBS. The mean additional improvement of 9.5 points (17% of the maximum ARAT score) on the ARAT is clinically meaningful and exceeds the previously established minimal clinically important difference (MCID) of 5.7 points (10% of the maximum ARAT score).^45^ The treatment benefit was observed directly after the two-week treatment period and until follow-up at 3 months after stroke. The therapeutic effect could not be established at 6 and 12 months post-stroke, possibly due to a plateau effect on the ARAT. A similar treatment effect was observed for the FMA arm score, which was also clinically meaningful,^37^ and the Barthel Index. However, other tests (NHPT, SULCS, JTT) in the activity domain did show a treatment benefit up to 12 months post-stroke. These tests are more sensitive to improvement in fine motor skills^46^ and manual dexterity.^41^ Patients in the sham cTBS group showed little improvement on these test after three months, while patients in the active cTBS group showed continued improvement of fine motor skills and manual dexterity up to 12 months post-stroke. We hypothesize that additional upper limb recovery in the first months after stroke, which may plateau out on commonly used upper limb motor scales, leads to more frequent use of the affected limb in more complex daily life activities, resulting in increased performance of fine motor skills and manual dexterity that persists over longer times.

Patients who received active cTBS showed more improvement on metrics of disability and dependency (mRS) and quality of life (SIS-UL) at 1 month post-treatment and at 3 months post-stroke, compared to patients who were treated with sham stimulation. The EuroQol-5D was a quality of life metric that did not show a treatment effect, presumably due to involvement of factors that are not directly related to an improvement in upper limb recovery, like pain and anxiety. In addition to a treatment effect on these metrics, the length of stay in the rehabilitation center, an indicator of independency, was 18 days shorter in patients treated with active cTBS compared to patients who received sham cTBS. These findings suggest that patients who received active cTBS were able to independently perform activities of daily living and participate in society at an earlier stage compared to patients who received sham cTBS.

Equally important to efficacy, the investigated cTBS treatment was safe and tolerable, as no serious adverse events were reported. Headache was more prevalent in the active cTBS group, but overall mild side-effects (headache/nausea) were uncommon (less than 4% of cases).

We did not detect an effect of active cTBS on contralesional M1 excitability, although a trend toward long-lasting (days) inhibition of the contralesional M1 could be observed. We speculate that active cTBS leads to a short-lasting reduction (< 2 hours) in contralesional M1 excitability, which has dissipated the next day, but which facilitates the effects of upper limb therapy directly following cTBS. Unfortunately, the acute effect of cTBS could not be measured as the cTBS treatments were directly followed by upper limb therapy. Our findings are in line with a recent meta-analysis that showed that contralesional inhibitory rTMS improves upper limb recovery on the FMA arm score with a similar mean difference of 9 points at 3 months post-stroke.^22^ Our study provides additional evidence that contralesional cTBS started within 3 weeks post-stroke effectively promotes upper limb recovery after stroke with a continued benefit until at least 12 months. Furthermore, we also observed a positive effect on scores of activity, disability and quality of life, which were not identified in aforementioned meta-analysis.^22^

Earlier meta-analyses of effects of rTMS treatment within the first months post-stroke were based (almost) exclusively on inhibition with conventional LF rTMS.^17,22^ A recently completed trial on contralesional LF rTMS in 77 patients did not find an effect on upper limb recovery.^47^ Continuous TBS has a substantially shorter treatment duration than LF rTMS, which increases patient comfort, makes it more suitable for use in time-constrained rehabilitation programs and potentially increases cost-effectiveness. An earlier study that assessed the impact of contralesional cTBS in the subacute post-stroke stage did not observe a clinical effect on motor function.^26^ This apparent discrepancy with our study may be explained by the smaller sample size (fourteen patients in the cTBS group), the later start of cTBS treatment with respect to stroke onset, the lower frequency of cTBS sessions (three sessions per week over three weeks), or the non-matching of upper limb physical therapy with cTBS sessions in that study.

A third of the screened patients did not meet the inclusion criteria because the treatment could not be started within 3 weeks after stroke onset. Extension of the treatment time window would have increased the number of eligible patients, but posed the risk of reducing the treatment efficacy, as a previous meta-analysis demonstrated efficacy for treatment only when started within the first month post-stroke.^22^

### Limitations

Our study had some limitations. First, the researchers were not blinded to the treatment allocation due to practical reasons concerning the sham condition, what could have introduced bias during the treatment period. In addition, while we performed double-blinded assessment of the primary outcome at three months, most secondary outcomes were assessed in a single-blind fashion. However, the considerably shorter length of stay in the rehabilitation center strongly suggests that the benefits observed on the other secondary outcomes are real.

Second, the sham condition did not consist of electrical stimulation to mask sensory sensations on the scalp evoked by peripheral nerve stimulation during TMS, and successful blinding of patients was not verified by asking patients which treatment group they believed they were assigned to. However, all patients were naive to cTBS treatment, which potentially reduces the bias introduced by limitations in the sham condition.

Third, we only measured the duration of upper limb therapy that patients received during the two-week treatment period and the week thereafter. However, it is unlikely that patients in both groups received different durations of upper limb therapy after this period, as all patients were part of the same treatment schedule. While upper limb therapy content was balanced between groups at onset of treatment, this may have changed based on additional improvement in upper limb function due to active cTBS, in accordance with the CARAS protocol. However, because both patients and therapists were blinded to treatment allocation, we consider it unlikely that patients in the active cTBS group received different upper limb therapy content unless the change in therapy was the direct result of improved motor function caused by active cTBS.

Fourth, we only scored the upper limb component of the FMA. Assessment of the lower limb component could have provided insights into the specificity of the treatment.

Fifth, the investigated sample consisted predominantly of males, which could limit generalizability of the results to females.

### Future research

The results in this single-center study are promising and provide a strong foundation for future multi-center trials to provide conclusive evidence on the efficacy of cTBS treatment in the promotion of upper limb recovery after stroke. These trials could tailor treatment to individual patients, as recent studies suggest that efficacy of contralesional cTBS may depend on stroke severity^48^ or stroke type.^49^

## CONCLUSION

In the present study, treatment with cTBS of the contralesional M1 combined with upper limb training, started within three weeks after stroke onset, improved upper limb motor recovery and led to better functional outcomes. Some treatment benefits persisted up to at least twelve months after stroke.

## Data Availability

Fully anonymous patient-level data can be made available on request.

## ACKNOWLEDGMENTS

We would like to thank all patients and investigators for their contribution to the B-STARS study.

## SOURCES OF FUNDING

This work was supported by the Netherlands Organization for Scientific Research (VICI 016.130.662) and in part by Brain Science Tools B.V. The funders had no role in study design, data collection, data analysis, data interpretation, or writing of the manuscript.

## DISCLOSURES

JV is a part-time employee of and SN is CEO and shareholder of Brain Science Tools B.V. Data analysis was performed during JV’s part-time employment at Brain Science Tools B.V. The other authors have no declarations of interest.

Fully anonymous patient-level data can be made available on request.

## SUPPLEMENTAL MATERIAL

Trial protocol Statistical analysis plan Table S1 and S2

## Notes

### Clinical Trial

The trial was registered at the international clinical trials registry platform (https://trialsearch.who.int/) with unique identifier: NTR6133.

### Clinical Protocols

https://bmjopen.bmj.com/content/7/8/e016566

### Author Declarations

The study was approved by the Medical Research Ethics Committee of the University Medical Center Utrecht.

